# Age Alters Integrated Cerebrovascular and Cardiovascular Dynamic Responses to Exercise: Insights from a Systems Modeling Approach

**DOI:** 10.1101/2025.05.02.25326897

**Authors:** Sandra A. Billinger, Eric D. Vidoni, Keshav Motwani, Bria L. Bartsch, Tyler Baldridge, Madeline Walker, Ali Shojaie

**Author notes:** **Corresponding Author:** Sandra A. Billinger, PhD, (913) 945 – 6685, 3901 Rainbow Blvd, Mail Stop 3051, Kansas City, KS 66160.

## Abstract

Understanding the dynamic interaction between cardiovascular and cerebrovascular systems during exercise is essential to evaluate the mechanisms supporting brain perfusion. This study examined age- and sex-specific differences in cardiovascular and cerebrovascular kinetics and used systems modeling to assess physiological coupling during moderate intensity exercise. We recruited adults to complete a single session of moderate intensity exercise on a recumbent stepper. Middle cerebral artery blood velocity (MCAv), mean arterial pressure (MAP), heart rate (HR), and end-tidal CO_2_ (P_ET_CO_2_) were continuously recorded. In 164 participants, the kinetic profiles were analyzed using mono-exponential modeling and functional data analysis. Granger causality within a subject-specific vector autoregression framework evaluated directional influence among physiological signals. Advancing age was associated with an attenuated dynamic response for MCAv, P_ET_CO_2_, and HR while MAP was elevated. Older adults exhibited significantly smaller MCAv amplitude and slower time constants than young and middle-aged groups. While sex did not influence overall MCAv, MAP, or HR kinetics, men had significantly higher P_ET_CO_2_ throughout exercise. Granger causality analysis revealed bidirectional coupling among MCAv, HR, MAP, and P_ET_CO_2_. Prior P_ET_CO_2_ levels significantly predicted MCAv while MAP had both short- and long-lag predictive effects on MCAv. MCAv also influenced subsequent changes in MAP and P_ET_CO_2_, indicating feedback regulation. P_ET_CO_2_ emerged as a dominant driver of MCAv, though systemic interactions reflect an integrated physiological network with multi-component feedback loops. This study advances understanding of cerebrovascular regulation and highlights the utility of systems modeling during exercise.

## INTRODUCTION

Understanding the dynamic interplay between cerebrovascular and cardiovascular systems during exercise is critical for elucidating the mechanisms that maintain cerebral blood flow and overall brain health. Exercise prompts a coordinated response involving cerebral blood flow regulation, partially driven by fluctuations in partial pressure of carbon dioxide (PaCO_2_), blood pressure (BP), cardiac output, neurogenic control, and heart rate (HR).(1) These interactions ensure adequate oxygen and nutrient delivery to meet the brain’s metabolic demands, which increase during varying exercise intensities (see in-depth review by Smith and Ainslie, 2017 (1)). Specifically, cerebral blood flow (CBF) “increases with exercise intensity up to approximately 60% of maximal effort,(2–4) after which cerebral blood flow may plateau during steady state exercise (5, 6) or decrease with near or maximal effort due to hyperventilation.”(7) These early studies shaped our understanding of cerebral blood flow responses across mild, moderate, and maximal exercise intensities during steady-state conditions.

Our previous work characterized the dynamic response of middle cerebral artery velocity (MCAv) during moderate intensity exercise, finding that MCAv increases exponentially after exercise onset in adults and plateaus during steady state exercise.(8) Indeed, exercise serves as a robust physiological stimulus that simultaneously engages multiple control mechanisms, including neural, metabolic, and vascular pathways.(1) Studying CBF responses to exercise offers critical insights into the resilience and adaptability of cerebrovascular control, with implications for both health and disease states. In addition, we showed older adults exhibit slower MCAv kinetics and reduced amplitude compared to young healthy adults,(8, 9) highlighting age-related impairments in cerebrovascular function and providing a framework for investigating cerebrovascular health. Despite prior work investigating MCAv responses to exercise, gaps persist in our comprehensive understanding of how age, sex, and the interactions among end tidal CO_2_ (P_ET_CO_2_), BP, and HR influence the cerebrovascular response across the lifespan. Resolution of the dynamic stimulus-response profile for P_ET_CO_2_, BP, and HR may provide a greater understanding of the underlying the physiological control processes than steady-state measurements alone.(10)

The purpose of this study was: 1) Characterize the interplay of cerebrovascular and cardiovascular regulation during exercise using our previous methods,(8) 2) Explore age- and sex-differences in the dynamic response profiles across cerebro- and cardiovascular measures, and 3) Apply an advanced modeling technique using Granger causality analysis to test whether past values of one physiologic signal contain information to predict future values of other physiologic signals. The integrated analysis of P_ET_CO_2_, BP, and HR provides novel potential insights into their collective and individual roles in driving MCAv responses. Our hypotheses were: 1) Following exercise onset, MCAv, HR, mean arterial pressure (MAP), and P_ET_CO_2_ would increase exponentially; 2) The kinetics profile would be altered with age such that the MCAv, HR, and P_ET_CO_2_ would be highest in the young adults, followed by middle age, and then older adults, whereas MAP would be lowest in the young adults, followed by middle age and highest in the older adults; 3) We would identify sex differences in MCAv, MAP, HR, and P_ET_CO_2_ responses during the transition from rest to exercise, with the magnitude being greater in men. Although P_ET_CO_2_ and MAP exhibit strong vasoactive properties on the cerebrovascular system, we hypothesized that P_ET_CO_2_ would have the greatest Granger causal influence on MCAv followed by MAP then HR.

## MATERIALS AND METHODS

We recruited a large, demographically diverse cohort spanning a broad adult age range and both sexes to enhance the generalizability of our findings in cardiovascular and cerebrovascular dynamics during exercise. All participants completed the study visit in a laboratory setting. Inclusion criteria were: 1) 20-85 years of age, 2) Willing to travel to our laboratory, and 3) Complete questionnaires and participate in a single bout of moderate intensity exercise. Participants were ineligible if any of these criteria were met: 1) Presence of absolute contraindications for exercise, as defined by the American College of Sports Medicine,(11) 2) Unable to perform alternating leg movements on the seated stepper exercise device, and 3) Required supplemental oxygen at rest or with exercise. The University of Kansas Medical Center Human Subjects Committee approved all study procedures, which complied with the Declaration of Helsinki. Institutionally approved written informed consent was obtained from each individual prior to study participation. Following consent, participants completed questionnaires regarding physical activity levels and aerobic fitness.(12, 13) Next, we screened participants for cardiovascular signs, symptoms, and diagnoses using the American College of Sports Medicine cardiac risk stratification.(11) Aligned with our previous work,(8, 9, 14) the study was completed in a dimly-lit, temperature-controlled environment. Prior to the visit, participants were asked to refrain from exercising for 24 hours, caffeine for a minimum of 8 hours, and food for 2 hours. Female participants were scheduled during the follicular phase of the menstrual cycle (days 1-7). (8, 9, 14)

The methods and experimental protocol used in this study have been published in detail in our seminal paper on the dynamics of middle cerebral artery blood velocity during moderate intensity exercise.(8) Briefly, participants first practiced the reciprocal motion of the recumbent stepper at the prescribed rate of 120 steps per minute. Moderate intensity was defined as 45–55% of HR reserve, calculated using the Karvonen formula. We calculated age-predicted maximum HR using 220 minus the age(11) or for those on beta blockers, we used 164-(0.7 × age).(15) We determined the target work rate by starting at 30 watts and increasing by 10 watts until the target HR zone of 45-55% HR reserve was reached and maintained. Participants were then instructed to stop exercise and rested until all physiologic signals returned to resting baseline levels.

### Equipment Instrumentation

While seated on the recumbent stepper, we instrumented participants with the following equipment: 1) Transcranial doppler ultrasound (Multigon Industries Inc., Yonkers, NY) for middle cerebral artery blood flow velocity (MCAv; cm/s), 2) 5-lead electrocardiogram (ECG, Cardiocard; Nasiff Associates) to monitor heart rate (bpm), 3) Finapres NOVA (Finapres Medical Systems, Amsterdam, the Netherlands) for beat-to-beat blood pressure (mmHg), and a 4) nasal cannula for PETCO_2_. We used the left MCA for data collection and analysis. If the left MCAv signal was unobtainable, we collected data from the right MCA. During the initial set up, participants sat quietly for 20 min before the exercise session began.

### Experimental Protocol

We recorded all physiological measures during 90 seconds of seated rest. At 20 seconds prior to the start of exercise, we provided standardized instructions that exercise would begin. To avoid Valsalva response and large fluctuations in the selected physiological measures, we used our standardized exercise initiation where participants began exercising at 60% of their target watts and increased the workload at 10-s intervals to reach the target watts 30 seconds into the exercise session.(8)

### Data Acquisition

Aligned with our prior work,(8, 9) all physiological variables were sampled at 500 Hz using an analog-to-digital unit (NI-USB-6212; National Instruments) and a custom-written MATLAB script (v2019a; TheMathworks Inc) for data acquisition and for data post-processing. To analyze, the data were divided by R-to-R cardiac interval. We calculated HR and P_ET_CO_2_ and the area under the curve was calculated for MCAv and MAP. All variables were then interpolated to 2.0 Hz using shape-preserving, piecewise cubic interpolation.

### Model Fitting

MCAv, HR, MAP, and P_ET_CO_2_ kinetics were modeled using the following mono-exponential equation, as previously published (8):

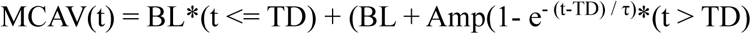

t indicates the variable value at any time point, BL, the baseline value before exercise onset, amp, the peak amplitude of the variable’s response above BL, TD, the time delay proceeding the variable’s exponential rise, and τ, the time-to-63% of steady-state.

### Functional Data Analysis: Sex and Age

To explore sex- and age differences in the dynamic response profiles across cerebro- and cardiovascular measures, we used a functional data analysis (FDA) approach,(16) as implemented in the “fda.usc” R package.(17) ‘Time was parameterized relative to stimulus onset (−89.5s to 360s) at 2Hz intervals.

### Systems modeling with Granger Causality Analysis

We investigated Granger causal effects for each physiological factor on all other measured physiological factors based on measurements in 0.5 second lags. Physiological data, including HR, P_ET_CO_2_, MAP, and MCAv, were included in a Vector Autoregression (VAR) model for each subject. The VAR model of order *p*, referred to as a VAR(p) model, for the *i*th subject, assumes the following structure: 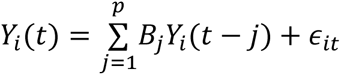

where *Y_i_*(*t*) is a vector containing the variables (MCAv, HR, MAP, P_ET_CO_2_) at the *t*th sample for the *i*th patient and *B*_*ji*_ is a coefficient matrix representing the effect of the variables at *j* samples prior on the immediate level (i.e. sample lag), and *∈*_*it*_ representing random noise. Separate models were fit for each subject with order between 1 and 50, and the median order minimizing the Bayesian Information Criteria (BIC) was selected as the final order, resulting in an estimated VAR(9) model. A VAR(9) model was subsequently fit for each subject separately, resulting in subject-specific estimates *B*_*ji*_ where *j* = 1, …, 9 and *i* = 1, …, *n* with *n* denoting the total number of subjects in the study. The residuals of the subject-specific fitted VAR(9) models were assessed to ensure stationarity, indicating adequate model fit.

### Statistical Analysis

We used one-way analyses of variance (ANOVA) with Tukey post-hoc adjustment to compare the dynamic response profile for our variables of interest across 1) sex and 2) age. The threshold for statistical significance was set at α < 0.05. Data are presented as mean ± standard deviation, unless otherwise specified. Analyses were performed with R Studio version 4.2.2.(18)

### Functional Data Analysis: Sex and Age Group

Group differences in modeled physiological responses (i.e., MCAv, MAP, HR, P_ET_CO_2_) over time were assessed using functional analysis of variance (FANOVA). For each signal, a global FANOVA was conducted to test the null hypothesis that all groups shared the same mean functional response over time. This test was based on a nonparametric bootstrapping procedure with 50 resamples. Pairwise contrasts were subsequently performed for all group combinations when the global test identified significant differences.

### Systems Modeling with Granger Causality Analysis

We tested whether there was any Granger causal effect for each physiological factor on others based on measurements in past samples. Specifically, assuming that the observed data were independent and identically distributed according to a common VAR(9) model across subjects, we tested whether the elements of *B*_*j*_ were nonzero and constructed confidence intervals for the elements of *B*_*j*_. To perform testing and constructed confidence intervals, we assumed that for each sample lag *j*, the estimates *B*_*ji*_ with *i* = 1, …, *n* were independent with an asymptotically normal distribution centered at *B*_*j*_ and the same variance. One-sample t-test using elements of the estimates *B*_*ji*_ with *i* = 1, …, *n* were employed to assess whether the effect was significantly different from zero. Bonferroni corrections were applied to control the family-wise error rate at 5% across the 144 tested coefficients, corresponding to 16 elements of each of the 9 coefficient matrices. Similarly, we constructed 95% Bonferroni-corrected confidence intervals for each element of the coefficient matrices *B*_*j*_ using the estimates *B*_*ji*_ with *i* = 1, …, *n*, controlling the family-wise mis-coverage rate at 5%.

Next, we tested whether the Granger causal effects differ by age and sex. Specifically, we assumed that the estimates *B*_*ji*_ with *i* = 1, …, *n* were independent with an asymptotically normal distribution centered at 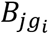 and the same variance, where *g*_*i*_ is the age group or sex of the *ii*th subject. The goal was to test if *B*_*j*,_*_young_* = *B*_*j*,_*_middle_* = *B*_*j*,_*_old_* and if *B*_*j*,_*_male_* = *B*_*j*,_*_female_*. A one-way ANOVA using elements of the estimates *B*_*ji*_ with *i* = 1, …, *n* with age group or sex as the factor was employed to test if there are differences in the elements of these coefficient matrices between age groups or sex. Once again, Bonferroni corrected p-values were used for the 144 tested coefficients for the age and sex comparisons separately.

## RESULTS

### Participant Demographics

We obtained data from 177 participants. However, 13 were excluded due to incomplete exercise protocols (n = 2), equipment failure (n = 3), or excessive signal noise precluding analysis (n = 8), resulting in 164 analyzable data sets (78 male, 86 female). Participant characteristics can be found in **Table 1**. The average participant age for this sample was 52.4 ± 21.3 years. This included 60 young adults (26.7 ± 5.1 years; 30 male), 26 middle-aged adults (53.9 ± 7.3 years; 13 male), and 78 older adults (71.7 ± 5.0 years; 35 male). Seven older adults reported use of beta-blocker. We analyzed the left MCAv in 95% of data sets (n = 155).

**Table 1.**
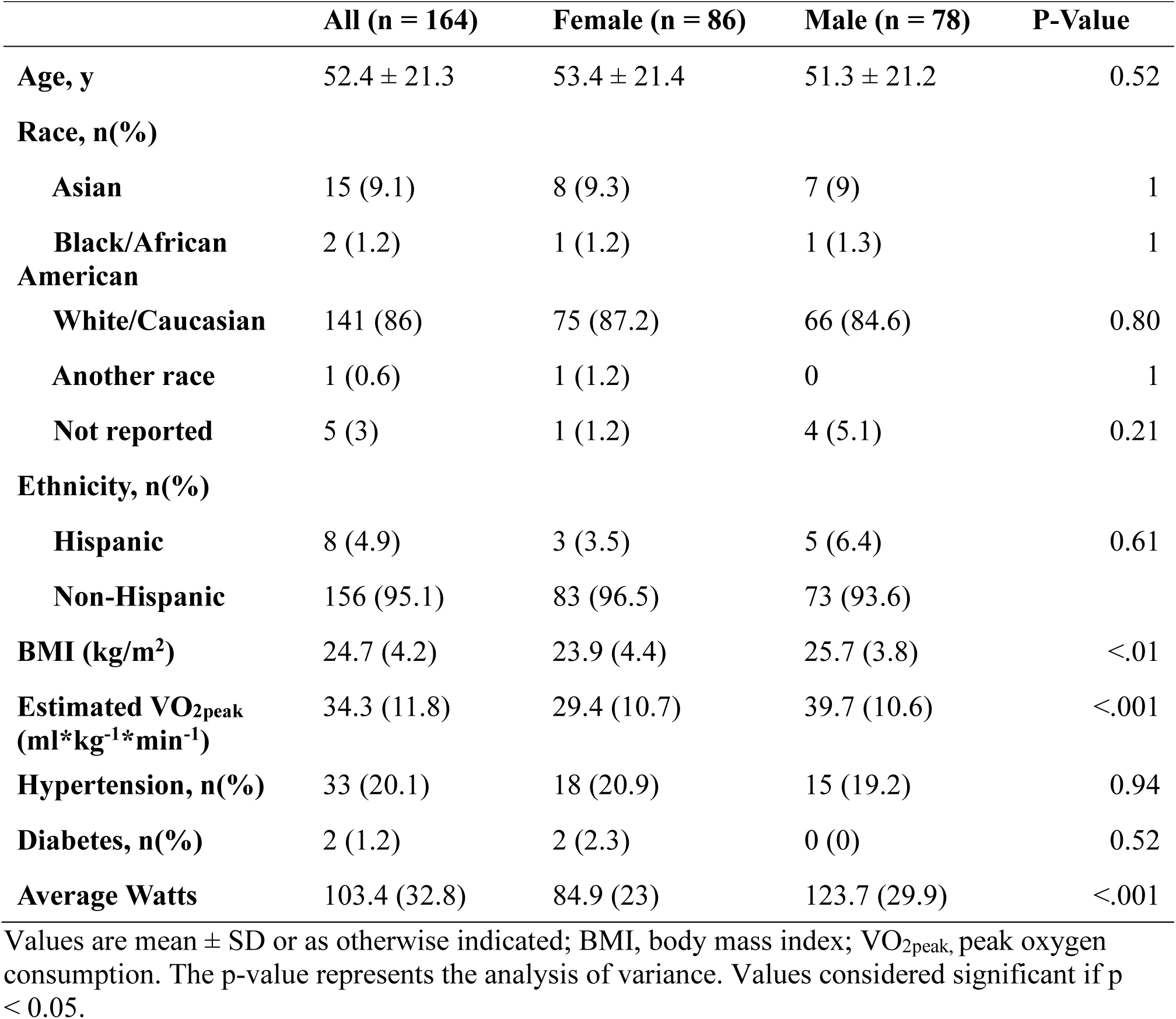
Participant Demographics.

### Dynamic Response Profile: Sex and Age

Overall, few sex differences were observed across cerebrovascular and cardiovascular measures. Females demonstrated significantly higher baseline MCAv compared to males (p = 0.002), and greater MAP time delay (p = 0.003). For P_ET_CO_2_, males had higher steady-state values (p = 0.001) and greater amplitude (p = 0.0006). No significant sex differences were found for heart rate. These differences, while statistically significant, were not consistent across all dynamic response profiles, indicating that sex-related effects were modest and measure-specific.

The data revealed significant age-related differences in the dynamic response profiles for MCAv, MAP, HR, and P_ET_CO_2_, during exercise (see **Tables 2-5** and **Figures 1-4**). MCAv baseline, amplitude, and steady-state values progressively decreased across age groups. In contrast, MAP increased with age, with higher baseline and steady-state values. P_ET_CO_2_ baseline and steady-state values declined with age, while baseline HR and steady-state values were highest in young adults and decreased with age. The data showed a negative HR time delay, suggesting a possible alerting or anticipatory response across all age groups, which may be associated to the timing of our instructions at 20 seconds prior to the start of exercise. Although our prior work reported a faster time delay in older adults (n = 14), those data were not significant.(9) Using a larger dataset, here, revealed a significantly faster MCAv time delay, in the older adults compared to the younger and middle-aged adults. While unexpected, we interpret our results in conjunction with previous research,(19) suggesting older adults may require additional cognitive and attentional resources allocated to the movement associated with the exercise bout. These findings were present in the older adult group (Figure 1b) and across both sexes (See Supplementary Figures 1 & 2). Overall, our findings highlight distinct, progressive age-related changes across the lifespan in the dynamic cerebrovascular and cardiovascular responses to exercise.

**Figure 1.**
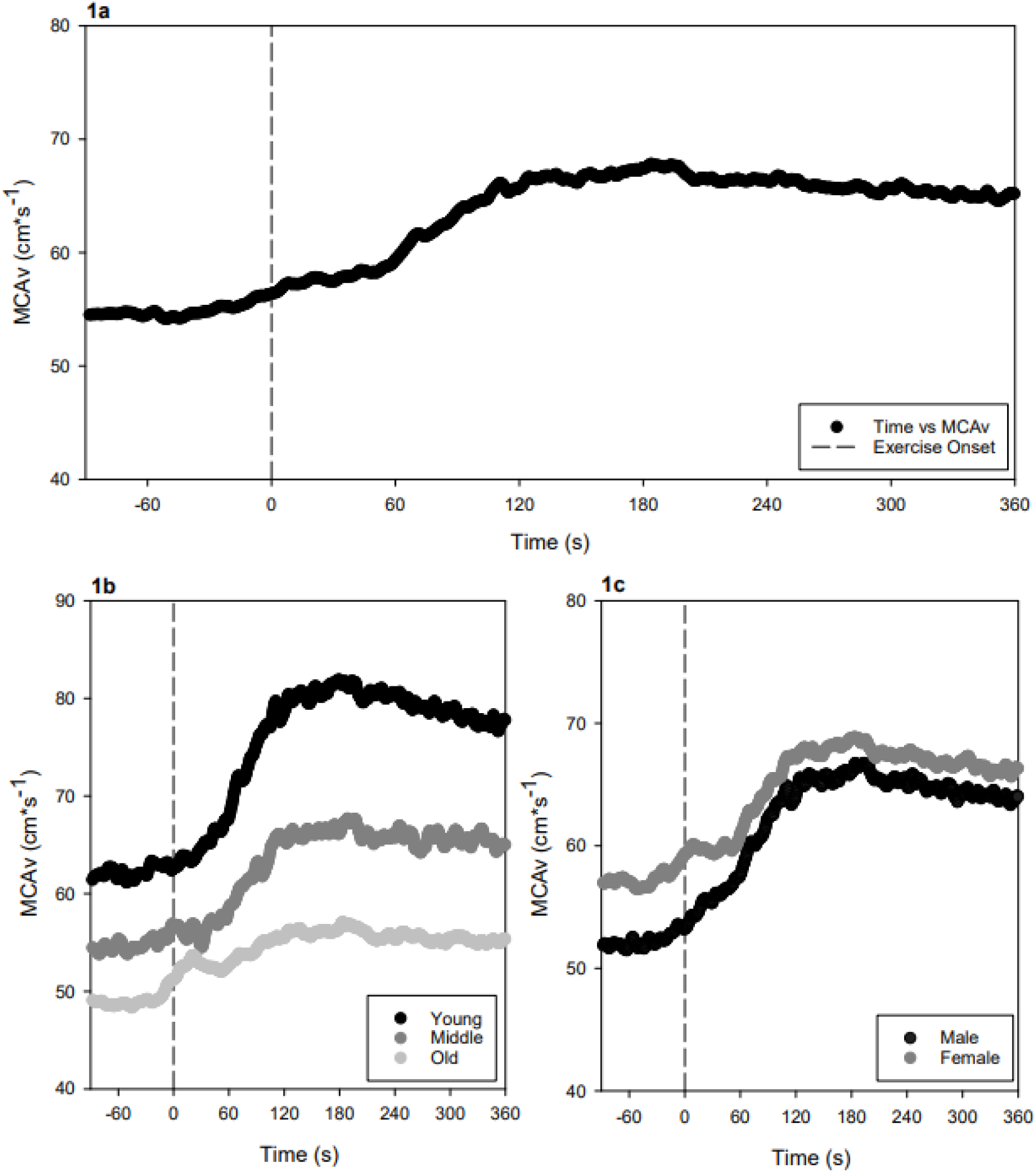
Middle cerebral artery blood velocity (MCAv) response to moderate-intensity exercise. Exercise onset (dashed line at 0 s). **1a** Group average MCAv (cm*s^-1^) across the entire sample (n = 164). **1b** MCAv response stratified by age group: young (black), middle-aged (gray), and older adults (light gray). **1c** MCAv response stratified by sex: males (black) and females (gray).

**Figure 2.**
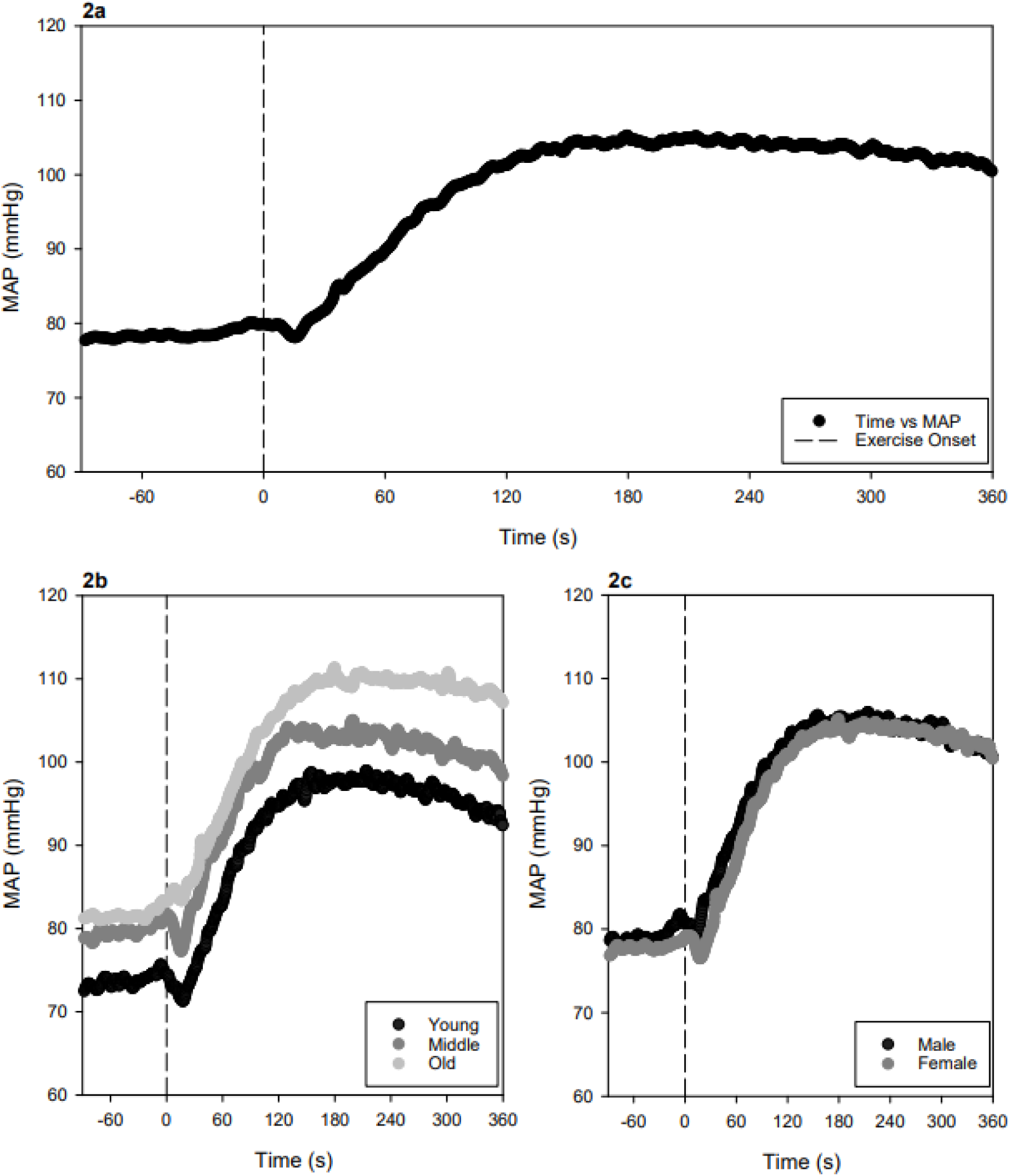
Mean arterial pressure (MAP) response to moderate-intensity exercise. Exercise onset (dashed line at 0 s). **1a** Group average MAP (millimeters of mercury; mmHg) across the entire sample (n = 164). **1b** MAP response stratified by age group: young (black), middle-aged (gray), and older adults (light gray). **1c** MAP response stratified by sex: males (black) and females (gray).

**Figure 3.**
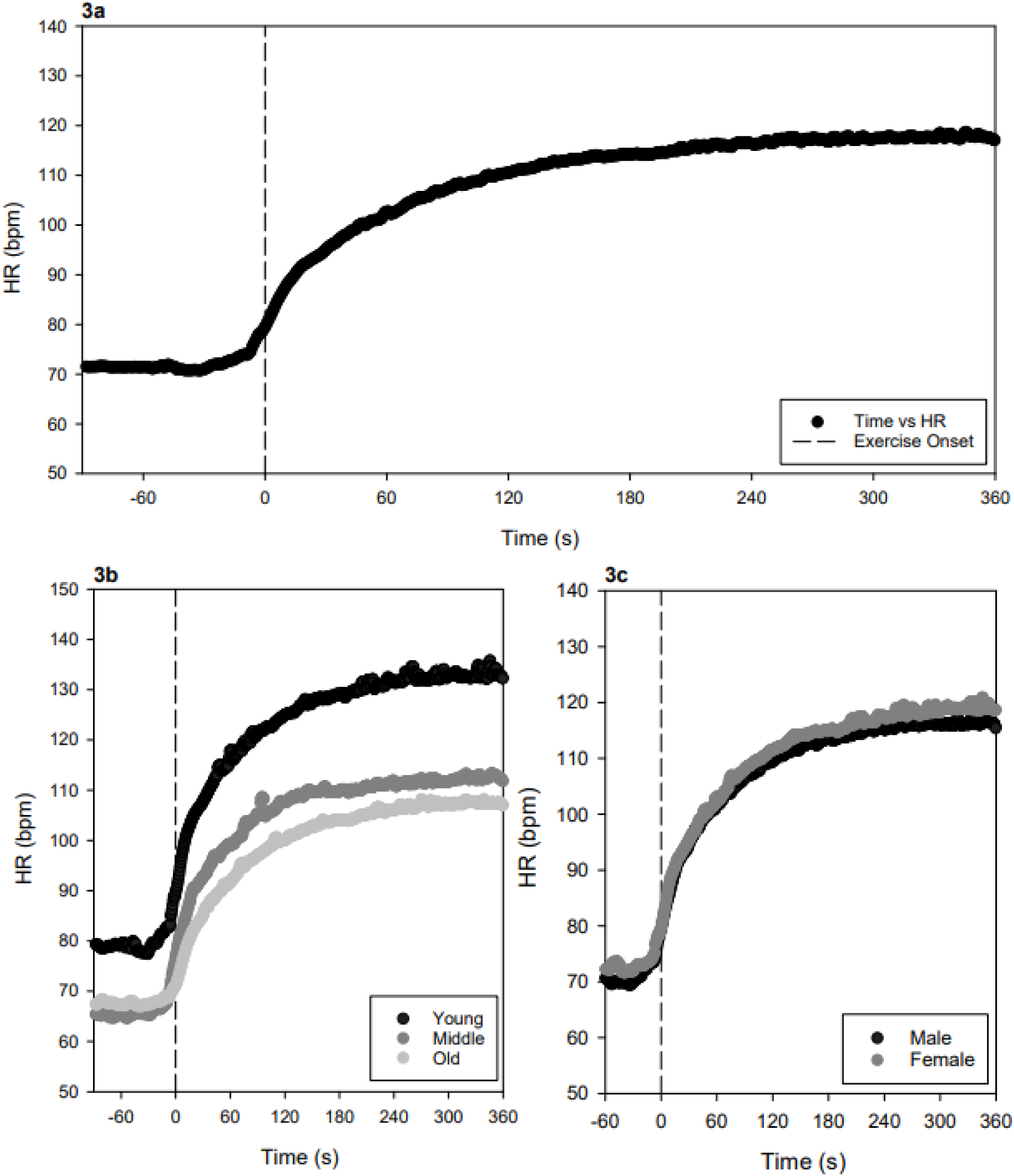
Heart rate (HR) response to moderate-intensity exercise. Exercise onset (dashed line at 0 s). **3a** Group average HR (beats per minute, bpm) across the entire sample (n = 164). **3b** HR response stratified by age group: young (black), middle-aged (gray), and older adults (light gray). **3c** HR response stratified by sex: males (black) and females (gray).

**Figure 4.**
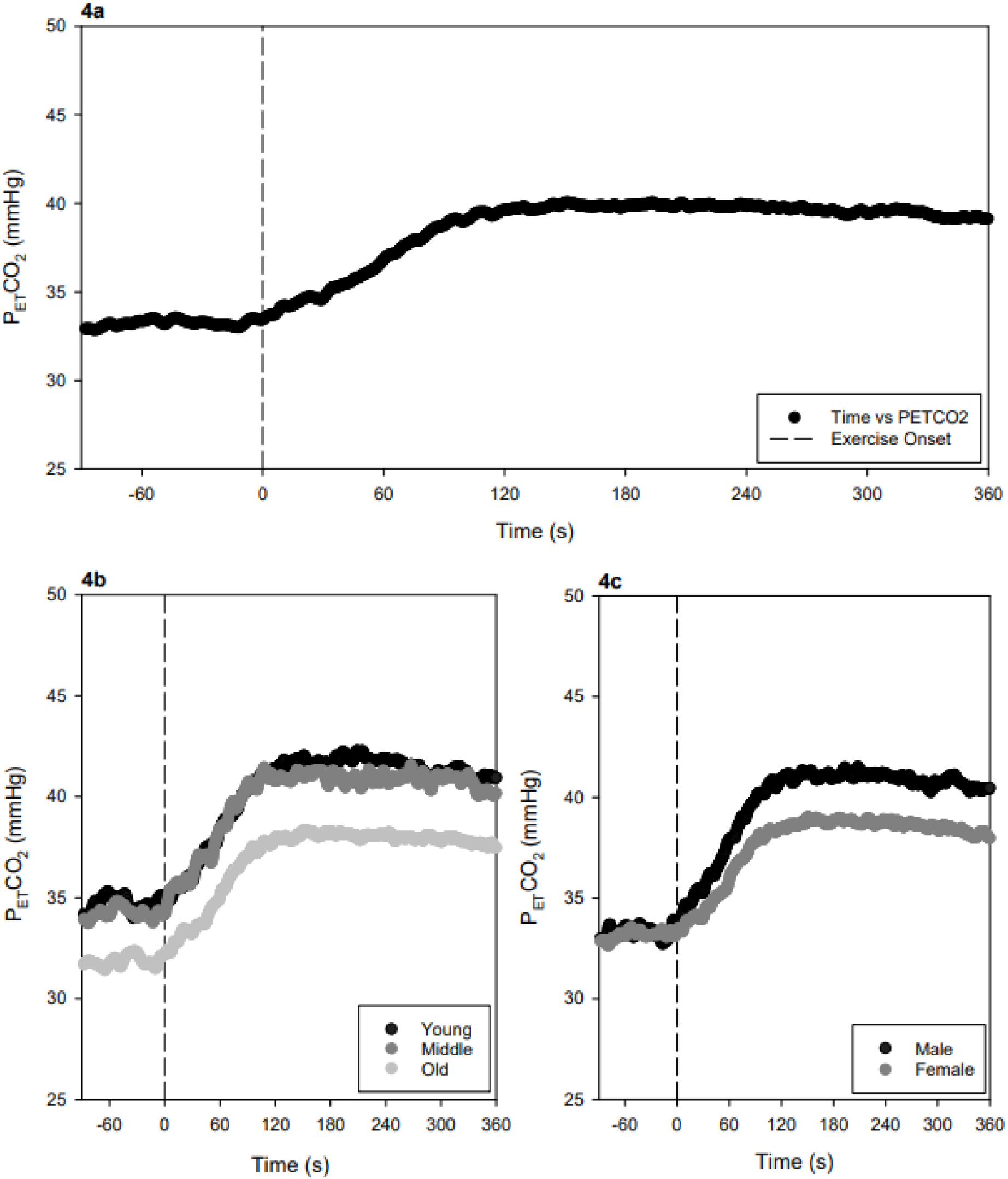
End tidal carbon dioxide (P_ET_CO_2_) response to moderate-intensity exercise. Exercise onset (dashed line at 0 s). **3a** Group average P_ET_CO_2_ (millimeters of mercury, mmHg) across the entire sample (n = 164). **3b** P_ET_CO_2_ response stratified by age group: young (black), middle-aged (gray), and older adults (light gray). **3c** P_ET_CO_2_ response stratified by sex: males (black) and females (gray).

**Table 2.** Middle Cerebral Artery Blood Flow Velocity Kinetics.

|  | Young | Middle-Aged | Older | P-Value |
| --- | --- | --- | --- | --- |
| **MCAv Baseline (cm\*s<sup>-1</sup>)** | 61.8 ± 10.6 | 54.8 ± 9.0 | 48.4 ± 10.6 | <0.001*^# |
| **MCAv Time Delay (s)** | 50.1 ± 23.3 | 60.2 ± 38.3 | 35.9 ± 42.4 | 0.01# |
| **MCAv τ (s)** | 29.0 ± 23.7 | 40.5 ± 44.4 | 36.6 ± 37.6 | 0.27 |
| **MCAv Amplitude (cm\*s<sup>-1</sup>)** | 17.4 ± 9.4 | 10.8 ± 5.9 | 6.2 ± 7.0 | <0.001*^# |
| **MCAv Steady State (cm\*s<sup>-1</sup>)** | 79.6 ± 15.6 | 65.2 ± 11.9 | 54.6 ± 12.5 | <0.001*^# |

Data are presented as mean ± standard deviation. MCAv, middle cerebral artery blood flow velocity; cm\*s<sup>-1</sup>, centimeters per second; s, seconds. The p-value represents the analysis of variance. Symbols indicate significant pairwise differences from Tukey post-hoc analysis: \*p ≤ 0.025 for young vs. older adults; ^p ≤ 0.025 for young vs. middle-aged adults; #p ≤ 0.025 for middle-aged vs. older adults.

**Table 3.** Mean Arterial Pressure Kinetics.

|  | Young | Middle-Aged | Older | P-Value |
| --- | --- | --- | --- | --- |
| **MAP Baseline (mmHg)** | 73.2 ± 13.6 | 77.3 ± 15.5 | 81.6 ± 13.6 | <0.01* |
| **MAP Time Delay (s)** | 44.4 ± 40.3 | 37.7 ± 19.9 | 23.7 ± 26.5 | <0.001* |
| **MAP $\tau$ (s)** | 36.6 ± 23.8 | 32.0 ± 20.1 | 55.7 ± 32.9 | <0.001*# |
| **MAP Amplitude (mmHg)** | 22.8 ± 12.8 | 24.0 ± 9.9 | 29.1 ± 9.7 | <0.01* |
| **MAP Steady State (mmHg)** | 97.1 ± 18.6 | 101.4 ± 17.8 | 109.7 ± 16.6 | <0.001* |

Data are presented as mean ± standard deviation. MAP, mean arterial pressure; mmHg, millimeters of mercury; s, seconds. The p-value represents the analysis of variance. Symbols indicate significant pairwise differences from Tukey post-hoc analysis: \* $p \leq 0.025$ for young vs. older adults; ^ $p \leq 0.025$ for young vs. middle-aged adults; # $p \leq 0.025$ for middle-aged vs. older adults.

**Table 4.** Heart Rate Kinetics.

|  | Young | Middle-Aged | Older | P-Value |
| --- | --- | --- | --- | --- |
| **HR Baseline (bpm)** | 78.7 ± 11.6 | 65.4 ± 10.4 | 67.5 ± 9.0 | <0.001*^ |
| **HR Time Delay (s)** | -16.1 ± 9.0 | -10.6 ± 7.0 | -11.8 ± 8.6 | <0.01^* |
| **HR $\tau$ (s)** | 70.8 ± 26.5 | 56.0 ± 22.5 | 83.6 ± 38.8 | <0.001# |
| **HR Amplitude (bpm)** | 54.8 ± 9.8 | 47.0 ± 7.2 | 41.3 ± 9.3# | <0.001*^# |
| **HR Steady State (bpm)** | 131.1 ± 10.6 | 110.8 ± 9.5 | 105.6 ± 10.3 | <0.001*^ |

Data are presented as mean ± standard deviation. bpm, beats per minute; s, seconds. The p-value represents the analysis of variance. Symbols indicate significant pairwise differences from Tukey post-hoc analysis: \* $p \leq 0.025$ for young vs. older adults; ^ $p \leq 0.025$ for young vs. middle-aged adults; # $p \leq 0.025$ for middle-aged vs. older adults.

**Table 5.** End-Tidal Carbon Dioxide Kinetics.

|  | Young | Middle-Aged | Older | P-Value |
| --- | --- | --- | --- | --- |
| **P<sub>ET</sub>CO<sub>2</sub> Baseline (mmHg)** | 34.6 ± 4.0 | 34.4 ± 2.8 | 31.6 ± 4.0 | <0.001*# |
| **P<sub>ET</sub>CO<sub>2</sub> Time Delay (s)** | 33.5 ± 53.0 | 42.5 ± 63.4 | 36.6 ± 47.3 | 0.77 |
| **P<sub>ET</sub>CO<sub>2</sub> τ (s)** | 41.3 ± 27.1 | 35.7 ± 21.1 | 46.6 ± 56.1 | 0.51 |
| **P<sub>ET</sub>CO<sub>2</sub> Amplitude (mmHg)** | 7.0 ± 5.0 | 6.7 ± 5.2 | 6.0 ± 4.4 | 0.46 |
| **P<sub>ET</sub>CO<sub>2</sub> Steady State (mmHg)** | 41.8 ± 5.2 | 41.0 ± 5.2 | 37.5 ± 4.7 | <0.001*# |

Data are presented as mean ± standard deviation. P<sub>ET</sub>CO<sub>2</sub>, end-tidal carbon dioxide; mmHg, millimeters of mercury; s, seconds. The p-value represents the analysis of variance. Symbols indicate significant pairwise differences from Tukey post-hoc analysis: \*p ≤ 0.025 for young vs. older adults; ^p ≤ 0.025 for young vs. middle-aged adults; #p ≤ 0.025 for middle-aged vs. older adults.

### Functional Data Analysis: Sex and Age Group

The global FANOVA also revealed significant sex-related differences in functional responses for P_ET_CO_2_ (p < 0.001), with men having generally higher P_ET_CO_2_ than women. But no differences were found for MCAv, MAP, or HR (p > 0.1). The global FANOVA also revealed significant age-related differences in the functional response in all physiologic signals (p < 0.01). Within MCAv, all three age groups differed significantly from each other (p < 0.001). For MAP, only the young age group was significantly lower than the older group (p ≤ 0.001). For HR, all age groups differed significantly from each other (p < 0.05). For P_ET_CO_2_, the young and middle age groups were significantly higher than the older group (p < 0.001).

### Systems Modeling with Granger Causality Analysis

The estimated systems model of the physiological data is shown in **Figure 5**. The results show pointwise confidence intervals and statistical significance for Granger causal effects of past physiologic signal values of one variable, e.g. P_ET_CO_2_, on another, e.g. MCAv, adjusting for past values of all other signals. The results corroborate the overall hypothesis that MCAv was both driven by and drives other physiological factors. That is, levels of other physiological factors at past timepoints were predictive of MCAv, and MCAv also predicted subsequent values of these physiological factors. For example, observed P_ET_CO_2_ at 2 sample lags (1 second prior) was significantly predictive of the immediate MCAv. Similarly, HR at the past 2-4 sample lags (1-2 seconds prior) was significantly predictive of immediate MCAv. MAP had both short and long-term effects on MCAv, with the MAP levels at the past 4 sample lags (2 seconds prior) and MAP levels at 8-9 lags (4-4.5 seconds prior) predicting immediate MCAv. Additionally, MCAv at 1 sample lag (0.5 seconds prior) was predictive of immediate P_ET_CO_2_ and MAP. Finally, the impact of HR on MAP followed a similar pattern as its impact on MCAv. Past values of each physiological factor were also highly predictive of current values of itself, indicating high levels of autocorrelation in each variable.

**Figure 5:**
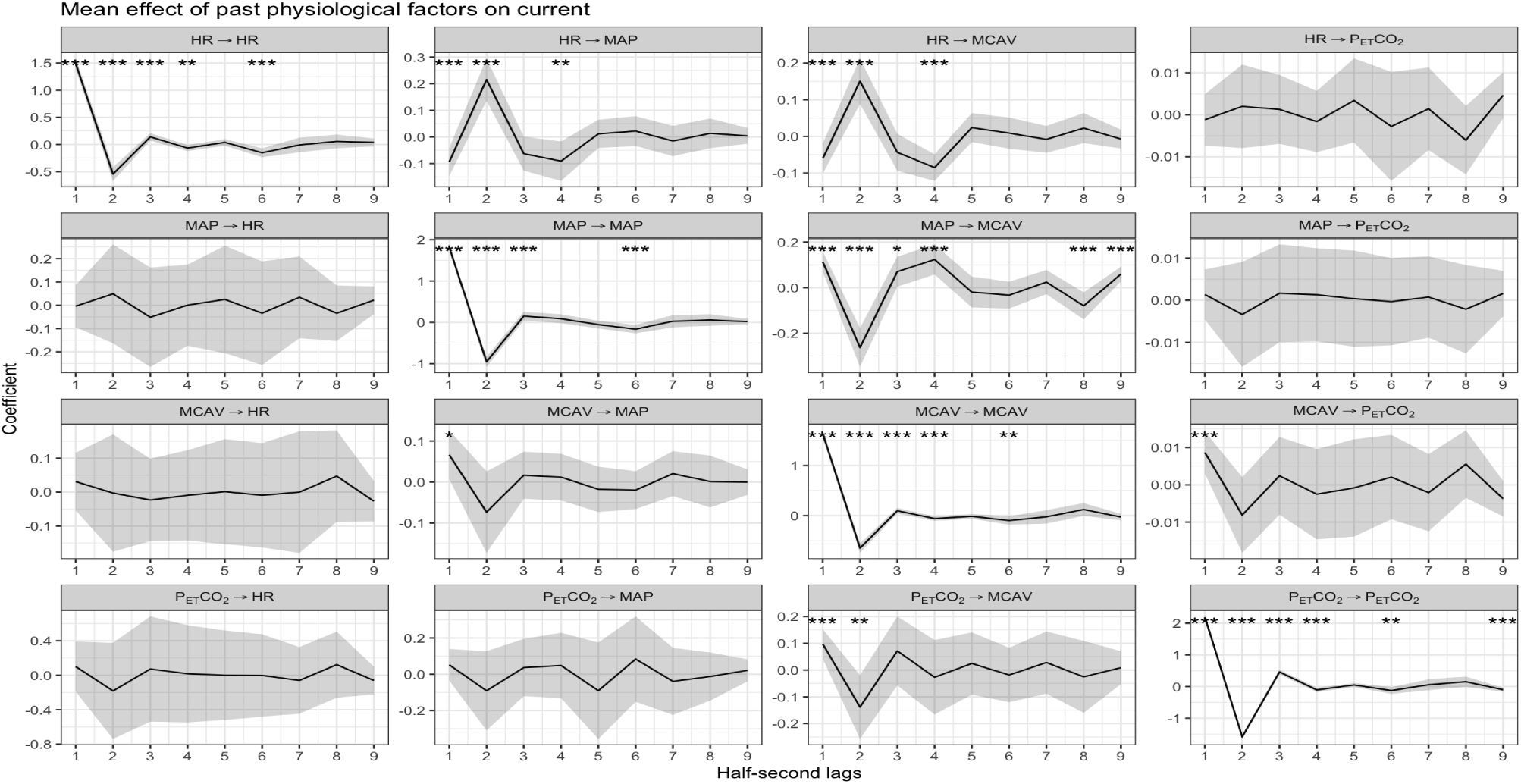
Estimated system VAR(9) model of physiological data. Each panel shows the estimated effect of one physiological factor from the past 9 lags (4.5 seconds) on the immediate level of another physiological factor (e.g. P_ET_CO_2_->HR indicates the effect of P_ET_CO_2_ level on HR). Bonferroni-corrected 95% confidence intervals of the estimates are plotted, as well as stars indicating the level of statistical significance testing whether the corresponding true effect is nonzero after applying Bonferroni correction across all 144 tests.

Next, we explored differences in the systems model of physiological data between young, middle-aged, and older participants in Figure 6. The results show participant-level estimates of the Granger causal effects, split by age group, with statistical significance for the difference of the effect of a past physiological factor on the current value between age groups. We observed many differences in how past values of a physiological factor impact the current value of that same factor (diagonal panels of **Figure 6**). For HR and MCAv, there are differences in both shorter- and longer-term effects of past values, whereas for MAP the differences are for medium-term effects, and the differences between age groups are restricted to longer-term effects for P_ET_CO_2_. We also observe differences between age groups in the effects across different physiological factors, for the effects of HR on MAP, HR on MCAv, and MAP on MCAv.

**Figure 6:**
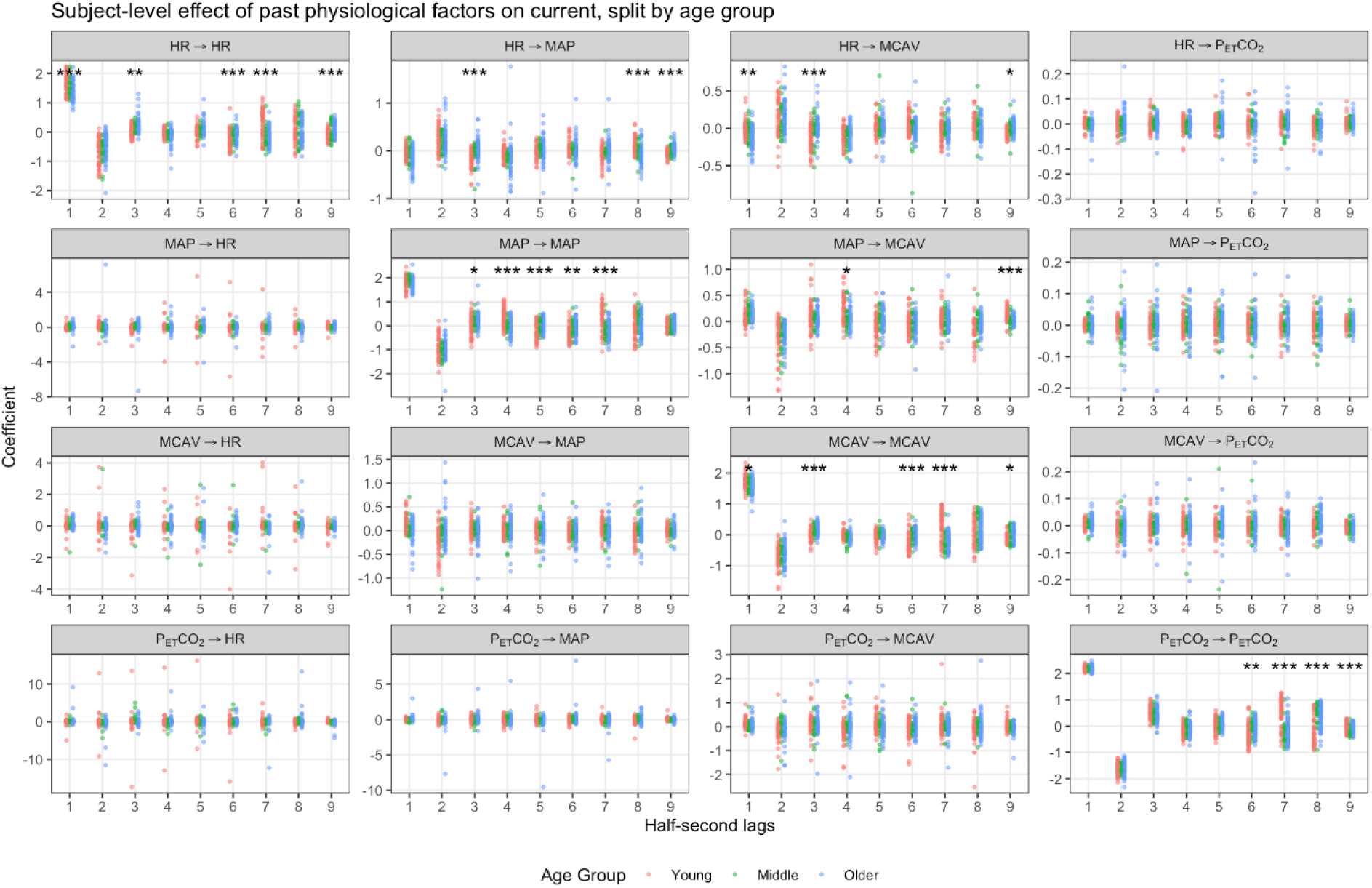
Estimated coefficients for VAR(9) model of physiological data for each subject, split by age. Each panel shows the estimated effect of one physiological factor from the past 9 lags (43.5 seconds) on the current level of another physiological factor (e.g. P_ET_CO_2_->HR indicates the effect of P_ET_CO_2_ level on HR), for each subject colored by their age. Stars indicate the level of statistical significance testing whether there was a difference in the effect between age groups (young, middle-age, and older) after applying a Bonferroni correction for 144 tests.

Finally, we examined differences in the systems model between male and female subjects. Controlling the family-wise error rate at 5% across all 144 comparisons, there was only one statistically significant difference, in the Granger causal effect of MAP on MCAv at a 2.5 second lag. While we cannot conclude there are no sex differences, qualitatively we observe many more and stronger differences between age groups compared to those between males and females.

## DISCUSSION

Our results are among the first empirical points of evidence for bidirectional feedback between MCAv and other systemic physiological signals within the framework of this methodology, something our prior work by Witte et al.(14) speculated on but could not formally demonstrate. Additionally, we(8, 14) and others (20–23) found weak or inconsistent associations between MCAv and systemic variables such as MAP, P_ET_CO_2_, and HR during various physiologic challenges. The present study supports and advances the cerebrovascular field by applying a systems modeling approach in a larger, more heterogeneous cohort. By modeling temporally lagged, directional influences, we demonstrate that P_ET_CO_2_, MAP, and HR each have significant predictive effects on MCAv during moderate intensity exercise, and in turn, MCAv exhibited smaller predictive effects on these variables. These findings extend prior observations and underscore the utility of advanced modeling in dynamic physiologic regulation during exercise not readily captured by traditional methods.

### Sex-Specific Exercise Responses

In the present study, our hypotheses for sex effects were not fully supported. We found minimal to no significant sex differences (p>0.10) in MCAv, MAP, and HR dynamic response profiles during moderate intensity exercise, with comparable time courses and amplitudes across males and females. These findings suggest that, at the group level, cerebrovascular and cardiovascular responses are broadly similar between sexes when exercising at moderate intensity. In contrast, prior work from our laboratory(9) identified a significantly slower MCAv response (longer time constant, τ) in older females compared to age-matched males and young females. However, that study included a small sample of older adults (7 men, 6 women), which may limit generalizability. During a graded exercise test involving young adults (n = 26), Ashley and colleagues(24) found no significant sex differences in the MCAv response and Murrell and colleagues report no sex-specific differences during submaximal steady state exercise.(25) The absence of widespread sex differences in our current, larger and more diverse cohort may reflect broader interindividual variability or the attenuation of age-specific sex effects in MCAv when examined across a wider population. The only variable with a sex-specific difference was P_ET_CO_2_, with males having a significantly higher P_ET_CO_2_ for amplitude and steady state exercise than females (p < 0.001). These findings for P_ET_CO_2_ corroborate prior work in healthy adults, which may be the result of higher breathing frequencies and smaller tidal volumes in females.(26)

### Age-Related Differences in Cerebrovascular and Cardiovascular Kinetics

In contrast to the minimal sex effects, age had a profound impact on hemodynamic responses. Our data revealed a clear, progressive attenuation of MCAv kinetics with advancing age. Older adults had a significantly lower baseline MCAv and a much smaller exercise-induced increase in MCAv compared to younger individuals. On average, baseline MCAv declined from ∼62 cm*s^-1^ in young adults to ∼48 cm*s^-1^ in older adults, and the amplitude of the MCAv rise was reduced by ∼65% in older adults relative to young (∼6 vs 17 cm*s^-1^). These findings corroborate earlier studies demonstrating age-related blunting of cerebrovascular responses. For example, our seminal(8) and subsequent works(9) showed that healthy older adults have a markedly lower MCAv at rest and a blunted rise in MCAv amplitude at exercise onset compared to young adults. These results support and extend our prior work that MCAv is progressively altered across the lifespan, with middle-aged responses intermediate between young and older. Likely mechanisms include structural and functional vascular changes such as arterial stiffening or endothelial dysfunction that could limit the capacity to augment cerebral blood flow in older adults.(27–29) We report that τ was not significantly different, suggesting similar rates of exponential rise across age groups once the response was initiated. We observed a significant age effect with a faster TD in the older adults. While this result was unexpected, we hypothesize this faster response was likely due to the timing of our verbal instructions and an anticipatory effect regarding movement planning.(19) Furthermore, these findings support prior work suggesting there may be “more to the story at exercise onset”(30, 31) that warrants further exploration, especially in older adults where movement planning may occur.

Age-related differences were not limited to the MCAv. We also found divergent patterns in systemic variables, with older adults having higher blood pressure and lower P_ET_CO_2_ and HR during exercise compared to young adults. Baseline and steady-state MAP increased with age (older > middle-aged > young), consistent with age-associated hypertension and arterial stiffness. (see in-depth review by Sun(32)) Despite higher MAP in older individuals, their MCAv response was smaller, which may support findings from previously published work that cerebral autoregulation and vascular tone adjustments in older adults may constrain flow despite elevated MAP during physiologic challenges.(9, 22, 33) In addition to higher MAP values in older adults, Fisher and colleagues,(22) reported decreased P_ET_CO_2_ at rest and during exercise. However, these differences were not statistically significant, likely due to a small sample size (n = 19). We observed age-related differences such that P_ET_CO_2_ declined with age with older adults demonstrating a lower P_ET_CO_2_ at rest and during exercise than young adults. This is an important finding as the cerebrovascular field has demonstrated carbon dioxide is a potent regulator of cerebral vessel tone(1) and dynamic changes in estimated partial pressure arterial CO_2_ tension are central to driving the MCAv rise during exercise.(10) Thus, the attenuated MCAv response observed in older adults may reflect a combined effect of reduced P_ET_CO_2_ and impaired pressure–flow regulation, both of which are essential for maintaining cerebral perfusion during exercise.

### Dynamic Interactions Between MCAv, MAP, HR, and P_ET_CO_2_

We selected Granger causality modeling to address the very limitations highlighted by Hegde and Hughson,(10) who noted that the cerebrovascular response to exercise is not a simple direct outcome of increased work rate but rather the result of multiple, interacting physiological signals contributing to cerebrovascular regulation. In this large heterogenous sample, Granger causality within a vector autoregressive framework allowed us to assess the directionality, timing, and interdependence among key signals such as MAP, P_ET_CO_2_, HR, and MCAv. The results revealed predictive, bidirectional relationships between MCAv and the systemic variables during exercise. Changes in P_ET_CO_2_ emerged as a significant driver of immediate changes in MCAv whereby elevated P_ET_CO_2_ levels in the prior half-second predict an increase in MCAv. We also found that HR changes preceded MCAv changes over short timescales such that an increased HR in the previous ∼1-2 seconds forecasted a higher immediate MCAv. Mean arterial pressure had both short- and longer-latency effects on MCAv, with MAP in the prior 2 and even ∼4-4.5 seconds showing predictive power for current MCAv. The shorter lag effect of MAP on MCAv may indicate immediate pressure-passive changes in cerebral perfusion,(10, 34) whereas the longer-lag effect could relate to slower regulatory adjustments. Importantly, MCAv itself influenced the other physiologic variables. For example, a higher MCAv at 0.5 seconds of lag significantly predicted higher subsequent P_ET_CO_2_ and MAP. Indeed, our finding that MCAv “Granger-causes” MAP is consistent with the concept of bidirectional interactions or feedback mechanism for baroreflex-mediated heart rate and vascular adjustments in response to changes in cerebral perfusion pressure.(35) Overall, the Granger causality analysis supports a view of the exercise response as a tightly integrated network rather than a one-way cascade. MCAv is both influenced by and influences systemic factors, indicating continual feedback mechanisms. This aligns with recent time-series analyses showing that the combination of dynamic changes in arterial pressure and CO_2_ can explain the MCAv response during exercise transitions, rather than any single factor acting in isolation.(10)

Building on these findings, we further examined how age and sex influence these dynamic relationships. By modeling temporally lagged, directional influences, we demonstrated that P_ET_CO_2_, MAP, and HR each have significant predictive effects on MCAv during moderate intensity exercise. We observed widespread age-related differences in both within-factor and cross-factor Granger causal effects, suggesting that the dynamics of cerebrovascular regulation and physiological factors are altered across the lifespan. In contrast, only one statistically significant sex-related difference was observed after adjusting for multiple comparisons, and while this does not mean that sex-related differences are not present, the data suggest that the age-related effects are much stronger, and thus age, may be a more prominent factor than sex in modulating the relationships between cerebrovascular regulation and physiological factors. Future work should build on the findings presented here by examining how aging affects the strength or timing of these coupling relationships.

While this study provides novel insights with a large, heterogenous sample across the adult lifespan, several limitations warrant consideration. First, our design was cross-sectional, comparing different age groups at a single time point. Cross-sectional comparisons cannot establish causality or the trajectory of changes within individuals over time. Therefore, longitudinal studies are needed to confirm age-related changes within the same individuals (27) and across chronic conditions. Second, we did not conduct exercise testing to determine maximum heart rate, and the work rate needed to exercise at 45-55% heart rate reserve. Instead, as we have published previously, we relied on age-predicted estimates to guide exercise intensity.(8, 9) Third, as we acknowledged previously,(8, 9) we used transcranial Doppler (TCD) ultrasound to index CBF. TCD measures blood velocity in the MCA, assuming the artery’s diameter remains constant even during exercise. A previous study in older adults using 4D Flow MRI reported no significant change in MCA diameter during an 8 mmHg increase in P_ET_CO_2_.(36) Although the average change in P_ET_CO_2_ in our study was less than 8 mmHg (**Table 5**), we were unable to directly measure MCA diameter during exercise. Therefore, blood velocity should not be assumed to represent absolute cerebral blood flow. Fourth, we recommend limiting participant instruction prior to exercise onset as this may alter the kinetics profile as observed by the negative HR TD and possibly the faster MCAv TD in the older adults. Finally, while highly informative, Granger causality does not provide causal interpretations and can only be interpreted as predictive linear associations within the VAR model. Therefore, the systems model might not describe the causal mechanism between the four physiological factors, particularly if key processes (i.e.: cerebral metabolism, hormones, cardiac output) that are in the true system are not one of the four observed factors.

### Clinical Relevance and Implications

Understanding age and sex differences in cerebrovascular and cardiovascular dynamics during exercise has important clinical implications for aging and brain health. Our results demonstrate that healthy older adults exhibit a markedly reduced MCAv response to moderate intensity exercise, which may reflect a decline in cerebrovascular reserve. Over time, reductions in cerebral perfusion reserve may increase vulnerability to neurodegeneration and cerebrovascular disease. Consistent with this, we have shown that healthy older adults with elevated beta amyloid, who are at an increased risk for Alzheimer’s disease, display reduced cerebrovascular responses during exercise.(37, 38) Similarly, individuals post-stroke exhibit diminished MCAv responses both during moderate intensity exercise compared to age- and sex-matched controls and early after stroke.(8, 39–41) Together, these findings highlight the importance of early detection of cerebrovascular dysfunction and underscore the need to examine dynamic physiological regulation using systems-level approaches. In the present study, Granger causality analysis revealed that P_ET_CO_2_, MAP, and HR each exert unique, time-dependent influences on MCAv during exercise, and that MCAv also exerts feedback effects on systemic signals. These findings reinforce the concept that cerebrovascular regulation is governed by an integrated physiological network rather than isolated mechanisms, and that disruptions in these interdependent systems with aging or disease may have broader implications for cerebral perfusion and brain health.

## CONCLUSION

This study provides new insights into the cerebrovascular and cardiovascular dynamic responses to moderate intensity exercise across the adult lifespan. Using both mono-exponential modeling and Granger causality analysis in a large, heterogeneous cohort, we show that aging is associated with attenuated MCAv response during exercise along with lower P_ET_CO_2_, and higher MAP, with minimal sex differences aside from higher P_ET_CO_2_ in men. Granger causality modeling revealed that P_ET_CO_2_, MAP, and HR each exert temporally lagged, predictive effects on MCAv, while MCAv also influenced systemic variables, indicating bidirectional, integrated regulation.

## DATA AVAILABILITY

The protocol and datasets generated and/or analyses are available from the corresponding author upon reasonable request.

---

**Supplemental Figure 1:**
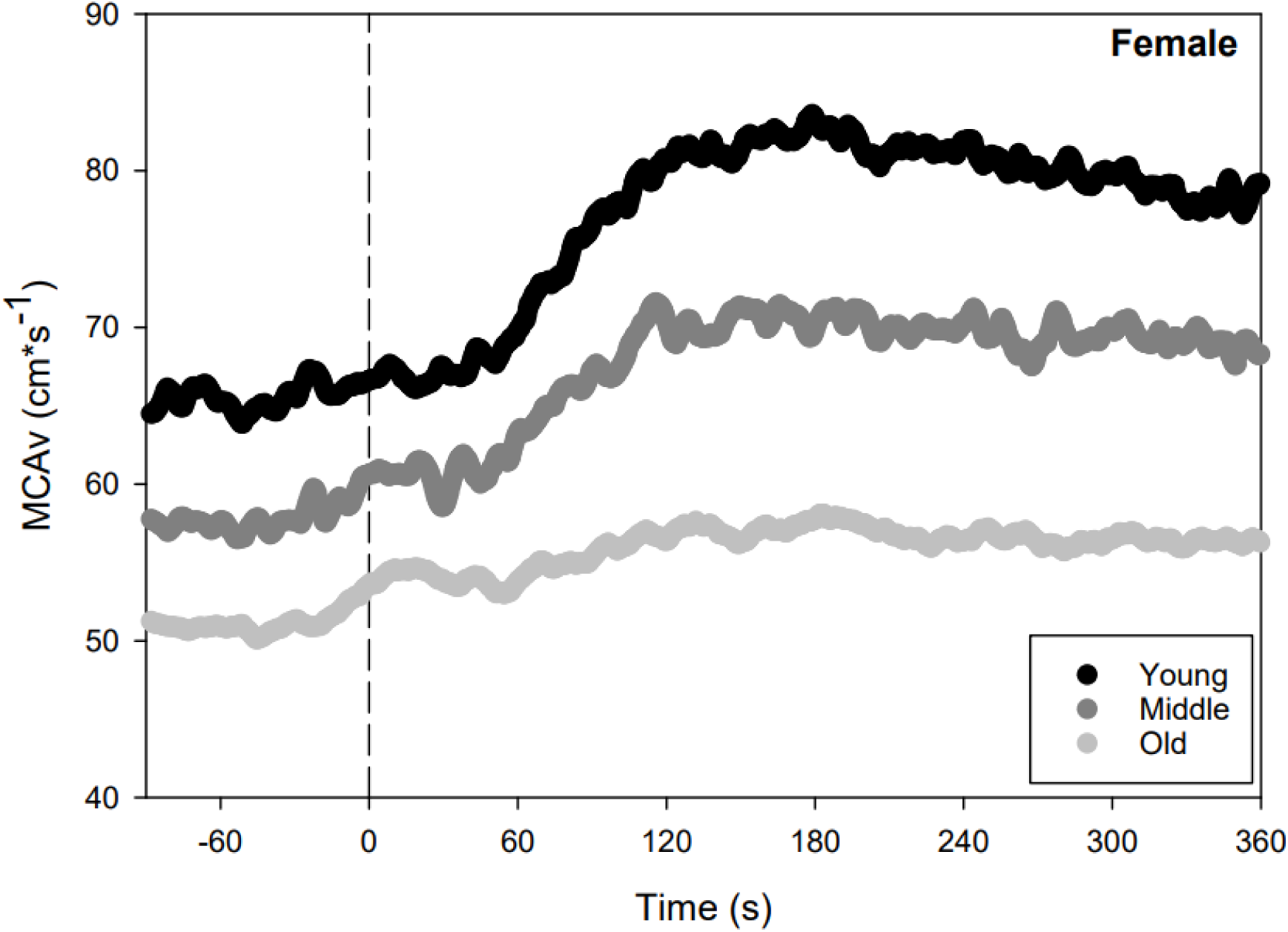
Middle cerebral artery blood velocity (MCAv) response to moderate-intensity exercise in females (n = 86). MCAv (cm*s^-1^) stratified by age group. young (black), middle-aged (gray), and older adults (light gray).

**Supplemental Figure 2:**
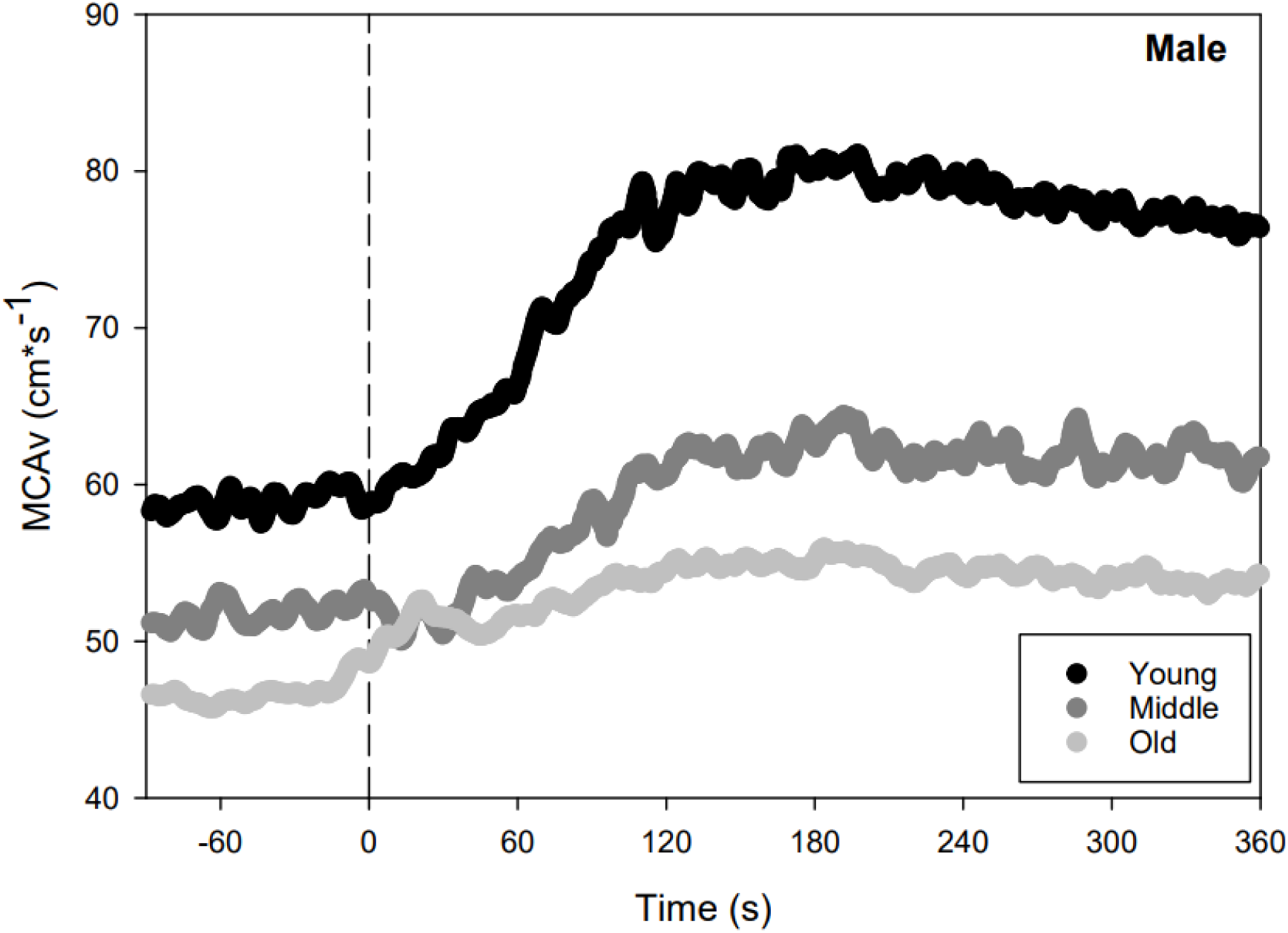
Middle cerebral artery blood velocity (MCAv) response to moderate-intensity exercise in males (n = 78). MCAv (cm*s^-1^) stratified by age group. young (black), middle-aged (gray), and older adults (light gray).

